# Acute respiratory distress syndrome and shunt detection with bubble studies: a systematic review and meta-analysis

**DOI:** 10.1101/2022.08.02.22278344

**Authors:** Jeffrey Odenbach, Sumeet Dhanoa, Meghan Sebastianski, Lazar Milovanovic, Andrea Robinson, Graham Mah, Oleksa G. Rewa, Sean M. Bagshaw, Brian Buchanan, Vincent I. Lau

## Abstract

**Objectives:** Acute respiratory distress syndrome (ARDS) is a life-threatening respiratory injury with multiple physiological sequalae. Shunting of deoxygenated blood through intra and extra-pulmonary shunts is one consequence that may complicate ARDS management. Therefore, we conducted a systematic review to determine the prevalence of sonographically detected shunt and its association with oxygenation and mortality in patients with ARDS.

**Data Sources:** We searched MEDLINE, EMBASE, Cochrane Library and DARE databases on March 26, 2021

**Study Selection:** Articles relating to respiratory failure and sonographic shunt detection.

**Data Extraction:** Articles were independently screened and extracted in duplicate. Data pertaining to study demographics and shunt detection were compiled for mortality and oxygenation outcomes. Risk of bias was appraised using the Joanna Briggs Institute and Newcastle-Ottawa Scale tools with evidence rating certainty using GRADE methodology.

**Data Synthesis:** From 4,617 citations, 10 observational studies met eligibility criteria. Sonographic detection of right-to-left shunt was present in 21.8% of patients (range:14.4-30.0%) amongst included studies using transthoracic, transesophageal and transcranial bubble Doppler sonography. Shunt prevalence may be associated with increased mortality (risk ratio: 1.22, 95% CI: 1.01-1.49, p=0.04, very low certainty evidence) with no difference in oxygenation as measured by P_a_O_2_:FiO_2_ ratio (mean difference -0.7, 95% CI: -18.6 to 17.2, p=0.94, very low certainty evidence).

**Conclusions:** Intra- and extra-pulmonary shunts are detected frequently in ARDS with ultrasound techniques. Shunts may increase mortality amongst patients with ARDS, but its association with oxygenation is uncertain. Future research should explore the role of shunt in ARDS, their association with mortality, and whether targeted precision medicine interventions can improve outcomes.

PROSPERO Registration Number: CRD42021245194 (March 26, 2021)

**Key Points:**

- **Question:** In adult critically ill ARDS patients, what is the prevalence of right-to-left shunts, and what are their effects on mortality and/or oxygenation?
- **Findings:** In this systematic review and meta-analysis, shunts be may prevalent in ∼1 in 5 ARDS patients. They may be associated with a statistically significant increase in mortality, with no difference in oxygenation parameters.
- **Meaning:** Intra- and extra-pulmonary shunts are detected frequently in ARDS with ultrasound techniques, and may increase mortality amongst patients with ARDS (although its association with oxygenation is uncertain).

## INTRODUCTION

Acute respiratory distress syndrome (ARDS) is a life-threatening lung injury that can occur following a variety of pulmonary insults including respiratory infection (e.g. bacteria or viral pneumonia, COVID-19) (1). Researchers describe several mechanisms that contribute to hypoxemia in ARDS (aside from parenchymal disease), including right-to-left shunts (2). Intra or extra-pulmonary shunting can occur from dysregulated pulmonary capillary deformation and/or intra-cardiac shunting via an intra-atrial septal defect, for example in acute cor pulmonale from elevated right-sided pressures during positive-pressure ventilation (3–5). This has become of increasing worldwide interest as the COVID-19 pandemic evolves and critical care units support patients with ARDS with refractory hypoxemia, where shunts have been hypothesized as a contributor to COVID mortality (6, 7).

The detection of right-to-left shunt has undergone significant transformation with the broader application and advancement of various sonographic modalities, with increased ease by point-of-care ultrasound (POCUS) providers (8, 9), and with backing from several echocardiography and ultrasound societies (10, 11). These include transthoracic echocardiography (TTE), transesophageal echocardiography (TEE) and transcranial doppler sonography (TCD), all of which can be used in conjunction with agitated saline bubble contrast administration to detect shunts (6, 7). Presence of agitated saline bubbles in the left-sided cardiac structures (TTE and TEE) or cerebral vasculature (TCD) indicates shunting of venous blood directly to the systemic circulation, bypassing pulmonary capillary vasculature (6, 7).

Therefore, we conducted a systematic review and meta-analysis to determine if the presence of shunt detection (using sonographic methods and contrast bubble studies) is associated with negative outcomes on oxygenation and mortality in ARDS

## MATERIALS AND METHODS

This systematic review was conducted in accordance with the PRISMA guidelines (12–14) and registered in advance with the international prospective registry of systematic reviews (PROSPERO CRD42021245194, registered March 26, 2021, search March 26, 2021). PRISMA checklist is shown in Supplemental Table 1.

### Search strategy

Searches were performed by a clinical librarian with experience in conducting electronic literature searches, with an additional librarian adjudicating using Peer Review Electronic Search Strategy criteria (11). We searched MEDLINE (via Ovid), EMBASE, Cochrane Library and DARE using combinations of keywords and, where appropriate, controlled vocabulary terms to identify articles pertaining to: acute respiratory distress syndrome, intracardiac or transpulmonary shunt, bubble contrast ultrasonography and related terms. Database search was executed on March 24, 2021, and followed guidelines described in the PRISMA statement (12–14). Detailed search strategy can be found in Appendix 1 of the supplemental material.

### Study selection and eligibility criteria

Articles were screened by title and abstract by two independent reviewers using the Covidence systematic review manager (www.covidence.org) (15) and selected for full-text review if identified as potentially relevant by one or more reviewers. Full-text review was done again in tandem by two independent reviewers and conflicts were resolved in discussion with a third reviewer. All article types were considered eligible that met the following criteria: (1) inclusion of adult patients with ARDS (including COVID-19) and (2) have undergone an agitated bubble saline sonographic study. Animal and pediatric articles were excluded, and no date or language restrictions were applied.

## Data abstraction and analysis

A pre-piloted data abstraction tables were created in Microsoft Excel version 14.0.6 (Redmond, WA, USA) and used by paired reviewers of included articles to extract study characteristics, patient demographic data, sonographic modality, shunt prevalence as well as oxygenation and mortality data, where available. We attempted to contact corresponding authors for retrieval of incomplete data where not directly published. Included data was manually verified for internal consistency between the paired reviewers to ensure final accuracy was maintained.

Continuous data were presented as means and standard deviations (SD), or medians and inter-quartile ranges (IQR), which will be compared (where appropriate) using a t-test or Wilcoxon rank sum test. Categorical variables and proportions will be compared using the Pearson’s χ^2^ or Fischer’s exact tests as appropriate.

Outcome data was compiled for meta-analysis using RevMan Cochrane software (https://revman.cochrane.org) (Copenhagen: The Nordic Cochrane Centre, Cochrane Collaboration 2014) version 5.4 software) (16, 17) and reported as relative risk (for mortality) and mean difference in P_a_O_2_:FiO_2_ ratio (for oxygenation) with significance set at 0.05. Confidence intervals (CIs) were reported for 95% CIs where applicable.

We used the method of DerSimonian and Laird to pool effect sizes for each outcome under a random-effects model for all outcomes of interest, with study weights measured using the inverse variance method (18). We presented the results as relative risk (RR) with 95% confidence intervals (CIs) for dichotomous outcomes. We presented the results as risk differences (RD) with 95% confidence intervals (CIs) for continuous outcomes.

Heterogeneity was assessed using the I^2^ statistic, the χ2 test for homogeneity (p <0.1 for significance of substantial heterogeneity), and visual inspection of the forest plots. We considered an I^2^ value greater than 50% indicative of substantial heterogeneity (16, 17). If significant unexplained heterogeneity existed, or if there is an insufficient number of studies for meta-analysis, we described data qualitatively. We assessed evidence of publication bias using funnel plots if there were 10 or more trials per outcome.

### Subgroup analyses

Potential and expected clinical sources of heterogeneity include different patient demographics, hospital characteristics and interventions strategies and follow-up. To explore significant heterogeneity, we planned the following pre-specified subgroup analyses, if a sufficient number of trials were available, (hypothesized direction of effect in parentheses):

- COVID ARDS vs. non-COVID ARDS studies (COVID studies would demonstrate worse shunt rates, hypoxemia and mortality compared to non-COVID ARDS).

### Risk of bias assessment and evidence grading recommendations

We assessed risk of bias using the Newcastle Ottawa Scale (NOS) and Joanna Briggs Institute (JBI) tools for observational cohort studies as described in our systematic review protocol (19, 20). Domains including selection (max score of 4), comparability (max score of 2) and exposure (max score of 3) were scored and reported as good/fair/poor based on domain scores of 3-4/2/0-1 (selection), 2/1/0 (comparability) and 3/2/0-1 (exposure) for the NOS tool.

We reported recommendations using the Grading of Recommendations Assessment, Development and Evaluation (GRADE) approach for mortality and oxygenation outcome data including risk of bias, inconsistency, indirectness, imprecision, and study source type to determine certainty of evidence to support the findings (21–23).

## RESULTS

### Study Characteristics

Our search yielded 4617 citations, with 51 relevant articles retrieved for full-text evaluation. Forty-one articles were excluded for: incorrect study design (25), incorrect patient population (1), missing outcome data (10), missing required intervention (3) and duplicate study (1). An additional article was excluded in the data extraction phase for incorrect patient setting (non-ICU) yielding a final inclusion of 10 eligible articles, where. The PRISMA flowchart is shown in Figure 1.

**Figure 1.**
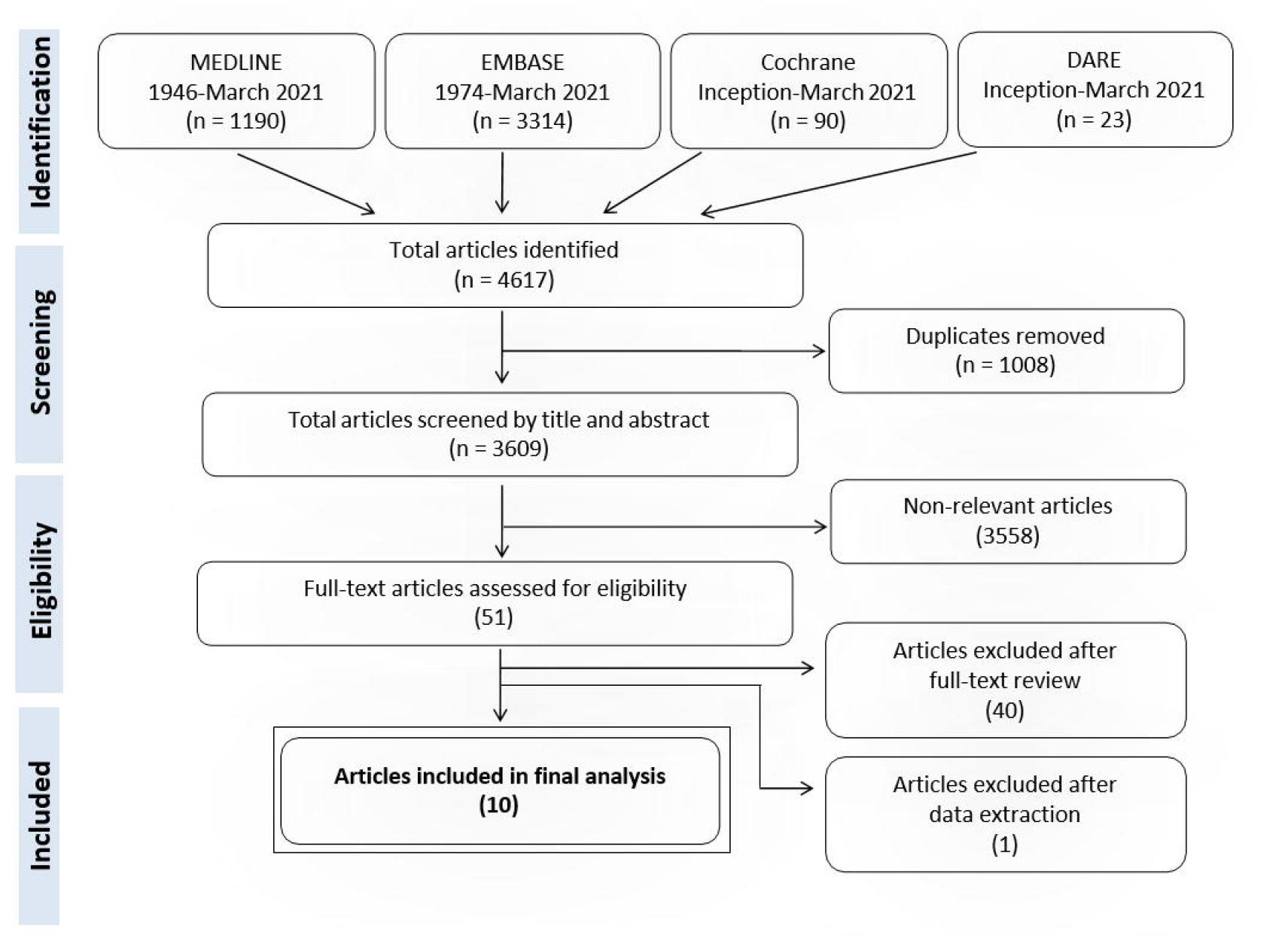
Flow diagram of systematic review selection criteria.

### Clinical outcomes

A total of 1,114 patients were pooled between 10 studies with an overall shunt prevalence of 21.8% as detected by various agitated saline bubble sonography modalities with individual studies ranging from 14.4-30.0% (2, 6, 7, 24–29) (2, 5–7, 21– 26). The majority of studies included ARDS secondary to infectious pneumonia, with 3 COVID-19 specific studies (6, 7, 24). Where reported, included studies demonstrated a male predominance (72.3%) and a mean age of 58.5 years. Remaining study demographics are shown in Table 1 (2, 5–7, 21–26).

Where ARDS mortality data was reported (n = 5 studies, 845 patients), the meta-analysis of pooled studies yielded 42.3% mortality (69/163 patients, 95% CI: 34.6-50.3%) for shunt presence compared to 32.0% mortality (218/682 patients, 95% CI: 28.5-35.6%) for shunt absence (risk difference: 10.3% [95% CI: 0.2-18.7%], relative risk [RR]: 1.22, [95% CI: 1.01-1.49], p=0.04, very low certainty). Forest plot for pooled mortality is shown in Figure 2 (2, 24–27).

**Table 1:** ARDS Shunt Articles Summary Statistics

| Reference: | N | Age (yrs) | % Male | SAPS II score | Primary diagnosis | Shunt Assessment modality | Overall Shunt prevalence |
| --- | --- | --- | --- | --- | --- | --- | --- |
| **Observational cohort (9)** |  |  |  |  |  |  |  |
| Boissier 2015 | 216 | 63 | 69.4 | 53 ± 25 | ARDS | TEE | 57/216 (26.4%) |
| Legras 2015 | 195 | 56 | NR | 46 ± 17 | PNA/ARDS | TEE | 28/195 (14.4%) |
| Lhéritier 2013 | 200 | 57 | 68.7 | 46 ± 17 | PNA/ARDS | TTE+TEE | 31/200 (15.5%) |
| Masi 2020 | 60 | 62 | 83.3 | NR | COVID-19 ARDS | TTE | 18/60 (30.0%) |
| Mekontso Dessap 2010 | 203 | 60 | 72.9 | 55 ± 18 | PNA/ARDS | TEE | 39/203 (19.2%) |
| Mekontso Dessap 2011 | 34 | 62 | 79.4 | 57 [40-72] | PNA/ARDS | TEE | 7/34 (20.6%) |
| Salazar-Orellana 2021 | 31 | 44 | 80.6 | NR | COVID-19 PNA | TCD | 7/31 (22.6%) |
| Vavlitou 2010 | 108 | 57 | 75.0 | NR | Respiratory failure | TEE | 30/108 (27.8%) |
| Védrinne 1995 | 49 | 53 | NR | NR | Respiratory failure | TEE | 11/49 (22.4%) |
| **Cross-sectional pilot study (1)** |  |  |  |  |  |  |  |
| Reynolds et al. 2020 | 18 | 59 | 61.1 | NR | COVID-19 ARDS | TCD | 15/18 (83.3%) |
| **WEIGHTED AVERAGES** |  | **58.5 ± 16.1** | **629/870 (72.3%)** | **50.4 ± 19.7** |  |  | **243/1114 (21.8%)** |

Data presented as mean (± standard deviation) or median [inter-quartile range] (% Male, SAPS II score). NR: Not reported, SAPS: Simplified Acute Physiology Score, ARDS: Acute Respiratory Distress Syndrome, PNA: Pneumonia, TTE: Transthoracic Echocardiography, TEE: Transesophageal Echocardiography, TCD: Transcranial Doppler.

**Figure 2.**
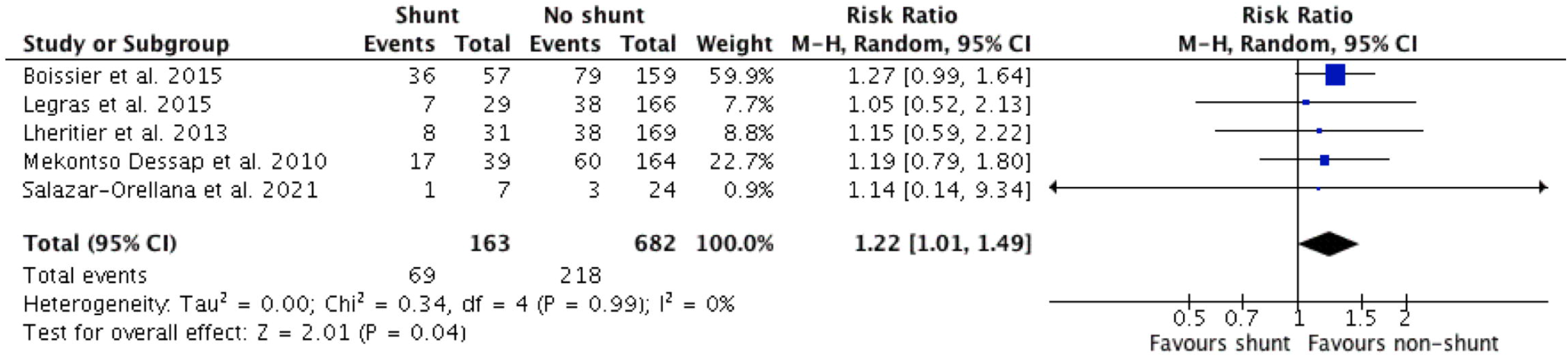
Composite meta-analysis of overall mortality

In contrast, oxygenation (as reported by P_a_O_2_:FiO_2_ [PF] ratio) was variable between reported studies (n = 5 studies, 700 patients). Shunt presence had a mean PF ratio of 123.8 ± 51.0 compared to 124.5 ± 46.3 for shunt absence. The mean PF ratio difference between groups is -0.7 (95% CI: -18.6 to 17.2, p=0.94, very low certainty). Forest plot for pooled PF ratio is shown in Figure 3 (2, 24, 25, 27, 28).

**Figure 3.**
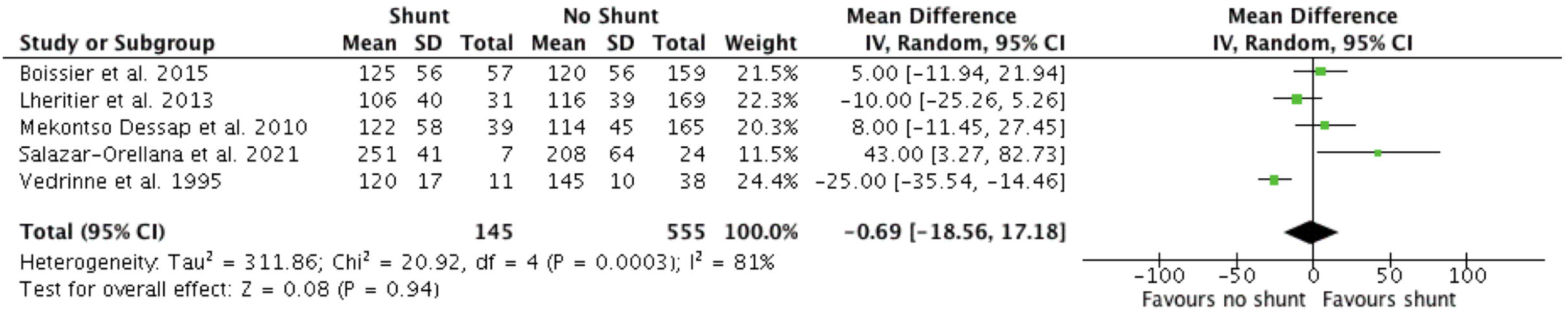
Composite meta-analysis of oveoxygenation (PaO_2_ / FiO_2_)

### Risk of bias, critical appraisal and publication bias

Risk of bias (RoB) was assessed using the Joanna-Briggs Institute (JBI) critical appraisal tool for cohort and case control studies (Supplemental Table 2) and the Newcastle-Ottawa Scale (NOS) tools (Supplemental Table 3). Overall assessment of “good” risk of bias was present in only 4 out of 10 (40%) of studies as assessed by the NOS. Similarly, binary deficiencies led to an overall appraisal of “include” in only 6 out of 10 (60%) of studies using the JBI tool.

The deficiencies leading to RoB assessment of “poor” (NOS) or “exclude” (JBI) were a primarily a result of unclear or absent description of comparator (shunt vs non-shunt, 2 studies) and absence of relevant oxygenation or mortality outcome data (4 studies).

Given there were less than 10 studies per outcome, no funnel plots were constructed for assessment of publication bias.

### Subgroup analyses

Pre-specified subgroup Forest plots for mortality and PF ratio are shown in Supplemental Figures 1 and 2.

For mortality (Supplemental Figure 1), there was a similar increased mortality for non-COVID (RR 1.22, 95% CI: 1.01-1.49; p=0.04, 4 studies) compared COVID-19 patients (RR 1.14, 95% CI: 0.14-9.34; p=0.90, 1 study), although not statistically significant.

For PF ratio (Supplemental Figure 2), there was equivocal effects for non-COVID (mean difference -6.7 (95% CI: -23.0 to 9.6; p=0.42, 4 studies) compared to COVID-19 patients 43.0 (95% CI: 3.3 to 82.7, p=0.03, 1 study), which was statistically significant in favour of better PF ratios with shunt presence.

## GRADE assessment

Composite study outcomes are summarized by GRADE assessment in Table 2. We identified an overall “very low” certainty of evidence for both outcomes of oxygenation and mortality.

**Table 2:**
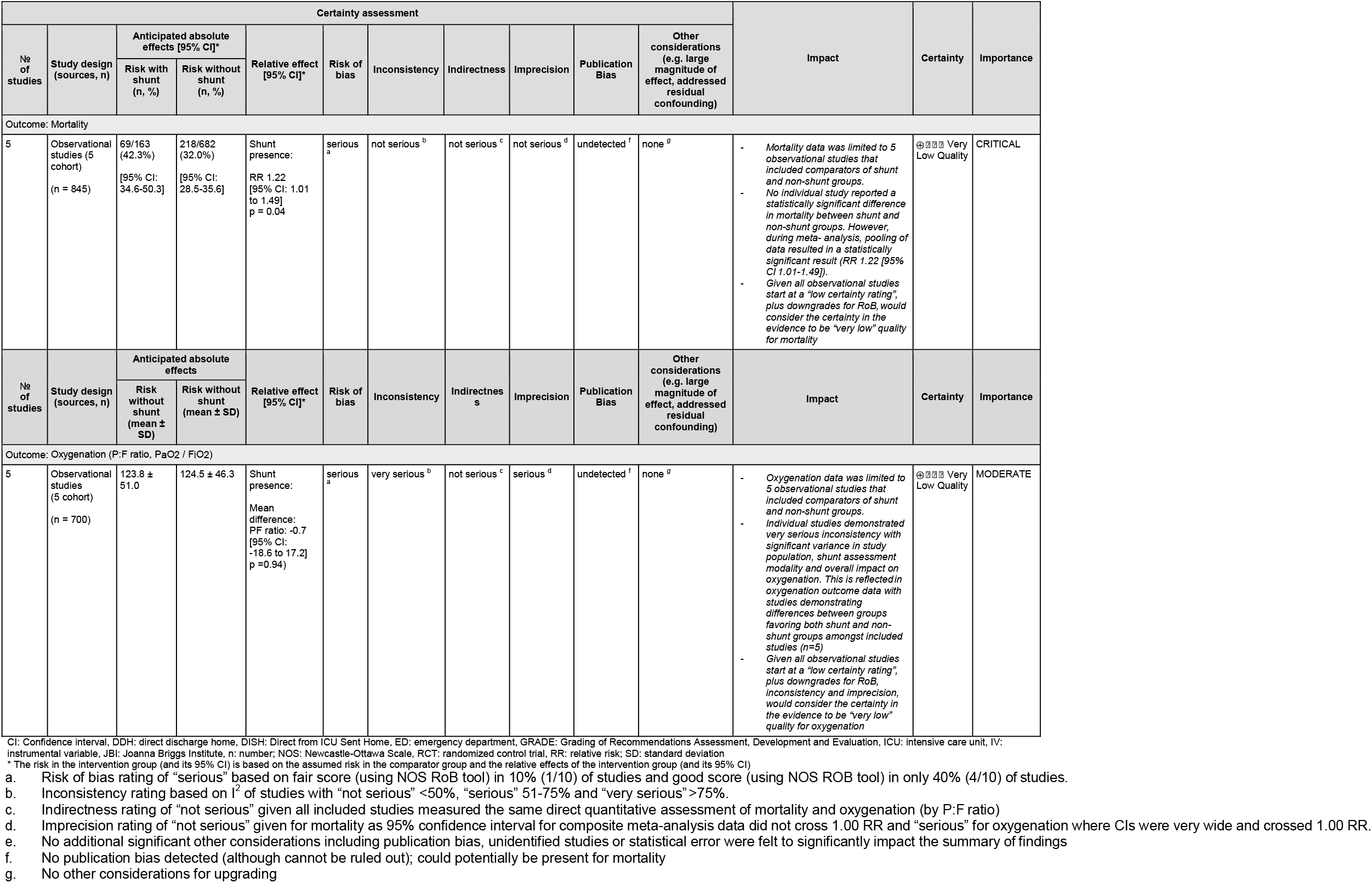
Grading of Recommendations Assessment, Development and Evaluation (GRADE) of ARDS Shunt Outcomes: mortality, oxygenation

For overall mortality, importance was deemed critical by direct clinical relevance. Inclusion of observational cohort studies limited certainty of evidence to “low” which was subsequently downgraded to “very low” due to serious risk of bias determined by the presence of a high proportion of studies receiving a score of “exclude” (4/10; 40%, JBI) or “poor” (5/10; 50%, NOS).

Similarly, composite meta-analysis data for oxygenation was downgraded to “very low” certainty of evidence based on high RoB scores detailed above, “very serious” allocation of inconsistency (studies with CI confined on either side of null effect) and imprecision (multiple studies with wide CIs).

## DISCUSSION

In this systematic review, we identified observational cohort studies describing techniques to detect intra and extra-pulmonary shunting and conducted a meta-analysis of mortality and oxygenation comparing patients with and without ARDS. Current literature demonstrates that right-to-left shunts are common, evident in approximately 1 in 5 ARDS patients (2, 5–7, 21–26). Our meta-analysis found increased mortality among critically ill patients with ARDS with sonographically detectable right-to-left shunt compared to no detectable shunt. It is unclear whether mortality is influenced by worse oxygenation levels, despite showing there may be no difference in oxygenation (with PF ratios reported) from this pooled meta-analysis. The evolving complexity of ARDS pathophysiology may include both direct and indirect effects on outcomes and physiology. Implementation of optimal therapeutic strategies and improving critical outcomes including mortality are goals that require ongoing advancement in the understanding of these mechanisms.

This study adds new knowledge regarding the prevalence of shunt in hypoxemic ARDS patients and its association on mortality, but not necessarily oxygenation alone. Shunt is on the differential diagnoses of refractory hypoxemia, with high alveolar-arterial gradients, but not necessarily always worked up with agitated saline contrast studies to confirm presence or absence of shunt. With approximately ∼22% shunt prevalence, this study demonstrates how many potential right-to-left shunts we might be missing if this entity is not investigated. What our systematic review failed to provide were further data on: 1) what type of shunt was present? (intra-cardiac versus intra-pulmonary); 2) what was done to address the presence of right-to-left shunt? (e.g. referral of patent foramen ovale closure, changes to ventilator management; 3) measurement of shunt fractions; 4) use of concomitant co-interventions (e.g. diuresis, inodilators or pulmonary vasodilators); 5) differences in duration of mechanical ventilation or supplemental oxygen used.

The finding of increased mortality in the shunt groups independent of oxygenation is difficult to conceptualize. However, limitations in oxygenation measurement using the PF ratio exist. The most accurate standardization uses a consistent FiO_2_ of 1.0 for PF ratio measurement (30), which is not routinely done in clinical practice. The PF ratios measured when patients are weaning from mechanical ventilation with lower oxygen requirements lead to non-standardized measures of PF ratio (31). Oxygenation is also incompletely evaluated by PF ratio, not necessarily accounting for applied positive end-expiratory pressure (PEEP) (31), which are not uniformly reported across all studies in this systematic review. Titration of PEEP with shunt presence may still lead to harms through various mechanisms, especially if the shunt is either intra-cardiac versus intra-pulmonary and is incorrectly treated with PEEP and mechanical ventilation.

There are many confounders which affect the outcomes of shunt presence, including: invasive mechanical ventilation parameters; heart-lung mechanics; RV dysfunction; baseline heart (e.g. congenital defects) and liver (e.g. hepatopulmonary syndrome in cirrhosis) comorbidities; temporal changes in physiology; non-standardized measurements of shunt; inter-rater reliability of diagnostics (3, 4). There are potential for all these biases due to differences in baseline risk, illness severity/acuity, amount and types of critical care support at the time of each study, and what diagnostics were available to the authors at the time of each study. Future research should seek to investigate potential confounders, minimize their impact, and standardize data collection.

Intra-cardiac shunts and intra-pulmonary are treated differently with mechanical ventilation and medications. For example, intra-cardiac shunts usually aim at lowering right-side heart pressures to prevent further shunting through the intra-atrial septum (e.g. patent foramen ovale [PFO], atrial septal defect [ASD]). Treatments may include: (1) diuresis; (2) pulmonary vasodilators (e.g. inhaled nitric oxide, epoprostenol) and/or inodilators (e.g. milrinone, dobutamine) through reducing RV afterload and improving RV function; (3) lowering ventilator settings (e.g. PEEP, plateau pressures); and, (4) closure of an intra-septal defects (PFO, ASD) to prevent further causing more R-L shunting (6, 7, 25, 32, 33). On the contrary, intra-pulmonary shunts are caused by pulmonary capillary vascular abnormalities, usually leading to abnormal vasodilation of pulmonary vessels. Therefore, mainstays of treatment may include: (1) avoiding pulmonary vasodilators (e.g. nitric oxide, epoprostenol) which would exacerbate hypoxemia; (2) careful titration of PEEP and ventilator settings to prevent dilation of pulmonary vessels, while preventing shunt from atelectasis from occurring; (3) reducing underlying cause of inflammation/infection, which led to the pulmonary vasodilation originally (e.g. corticosteroids in COVID-19 shunt) (7, 24, 34). Given these nuances, optimal conservative, non-procedural management of intra-cardiac vs. intra-pulmonary shunts is required. It is important that future work should focus not only on diagnosing the presence of R-L shunts, but also reporting what type (intra-cardiac vs. intra-pulmonary). This is to help tailor the correct ventilation and medical treatments individually for each ARDS patient, allowing for precision medicine. Future work should focus on determining if diagnosis of either intra-cardiac vs. intra-pulmonary shunt is important to distinguish, and if certain strategies to deal with each entity can improve outcomes.

The strengths of our SR include a comprehensive search strategy and a rigorous process for study selection and data abstraction based on an *a priori* protocol, with due consideration to study quality, risk of bias and overall certainty of the evidence using GRADE alongside our meta-analysis methodology.

This SR also has several limitations, most of which relate to limitations of the primary studies analyzed. GRADE certainty of evidence was very low for all outcomes, driven primarily by majority of studies observational in nature, without adjustment for baseline characteristics and illness severity) alongside small sample sizes. With significant variability in study design and focus, definitions of shunt were also variable between included studies. This ranged from the sonographic detection of right-to-left bubbles quantitatively from any number of detected bubbles by TCD at any time (6, 24) to at least 10 bubbles at any time by TEE (27) to any number of bubbles within three cardiac cycles (2). Although our composite data and meta-analysis support the finding of increased mortality associated with the sonographic detection of shunt, a causal relationship is difficult to rationalize and further research focusing on the significance of right-to-left shunt in ARDS is required. The level of data provided in these studies were lower, with many not measuring: shunt fractions, use of inodilators, or pulmonary vasodilators. Even differences in duration of IMV or total duration of supplemental oxygen use were not routinely measured. Lastly, there were no pre-specified ARDS subgroups of note, specifically based on ARDS severity and etiology. There is also substantial heterogeneity among ARDS patients with respect to how much shunts contribute to their illness (e.g. COVID deaths differed during different waves of the pandemic).

Our group has aimed to address this question further by conducting an observational cohort study using of TTE, TEE and TCD in the detection of shunt in a series of critically ill patients with SARS-CoV-2 (COVID-Shunt Study).

## Conclusion

The detection of intra and extra-pulmonary shunt in ARDS using ultrasonography is relatively common in critically ill patients with ARDS. There may be increased mortality among patients with ARDS and evidence of shunt (very low certainty). However, shunt prevalence may have uncertain direct physiologic impacts on oxygenation (very low certainty).

## Data Availability

All data produced in the present work are contained in the manuscript

## Acknowledgements

We are grateful to Diane Keto-Lambert (Faculty of Medicine and Dentistry, University of Alberta) for her assistance with the SR search, as well as during the PRESS process with Doug Salzwedel.

## Author Contributions

Jeff Odenbach, Vincent Lau, Sumeet Dhanoa, Lazar Milovanovic, Andrea Robinson, Oleksa Rewa, Sean M. Bagshaw and Brian Buchanan have: (1) made substantial contributions to conception and design, acquisition of data, analysis and interpretation of data; (2) drafted the submitted article and revised it critically for important intellectual content, and (3) provided final approval of the version to be published.

Conception: Lau, Odenbach, Buchanan Background: Lau, Odenbach, Mah, Buchanan

Design: Lau, Odenbach, Bagshaw, Rewa, Buchanan, Mah

Acquisition of data: Lau, Odenbach, Dhonoa, Milovanovic, Robinson, Mah

Analysis of data: Lau, Odenbach Dhanoa, Rewa, Bagshaw, Milovanovic, Robinson, Buchanan

Drafting the manuscript: Lau, Odenbach, Dhanoa, Rewa, Bagshaw, Milovanovic, Robinson, Buchanan

Revising the manuscript: Lau, Odenbach, Dhanoa, Rewa, Bagshaw, Milovanovic, Robinson, Buchanan

## Competing Interests and Funding

None of the authors disclose any competing interests. There was no funding for this research. Dr. Bagshaw is supported by a Canada Research Chair in Critical Care Outcomes and Systems Evaluation.

## Patient and public statement

Patients or the public were not involved in the design, or conduct, or reporting, or dissemination plans of our research.

## Tables

Table 1: ARDS Shunt Articles Summary Statistics

Table 2: GRADE Summary of Findings

## Figures

Figure 1: ARDS Shunt PRISMA Flowchart

Figure 2: Forest plot mortality for ARDS shunt vs. non-shunt patients

Figure 3: Forest plot oxygenation for ARDS shunt vs. non-shunt patients

## Notes

### Competing Interest Statement

None of the authors disclose any competing interests. Dr. Bagshaw is supported by a Canada Research Chair in Critical Care Outcomes and Systems Evaluation.

