## Supplemental Appendix 1 for "Acute respiratory distress syndrome and shunt detection with bubble studies: a systematic review and meta-analysis"

**Search Log for: COVID-19/ARDS and Shunts**

FINALIZED: 26 Mar 2021

**Electronic Databases Searches**

| **Import #** | **Database Name*** | **Search Interface** | **Database Dates** | **Date of Search**  **MM/DD/YYYY** | **Initial Count** | **Post De-dupe** |
| --- | --- | --- | --- | --- | --- | --- |
| 1 | Medline | Ovid | 1946 to Present | 03/24/2021 | 1190 | 1187 |
| 2 | Embase | Ovid | 1974 to Present | 03/24/2021 | 3314 | 2381 |
| 3 | Cochrane | Wiley | Inception – Present | 03/24/2021 | 90 | 22 |
| 4 | DARE | NHS | Inception - Present | 03/24/2021 | 23 | 23 |
| **Total Database Search Results**: | | | | | **4617** | **3613*** |

*An additional 5 duplicates were removed by Covidence for a total of 3608

**Location of search result folder and file names:** **Z:\KS Projects\Lau - ARDS Shunts**

**Librarian(s)/researcher(s) conducting search strategy** ((initials) name, degree(s))**:**

(DKL) Diana Keto-Lambert, MLIS; Peer review by Doug Salzwedel, MLIS

Appendix: Search Strategies

Database(s): **Ovid MEDLINE(R) ALL** 1946 to March 26, 2021

Search Title: ARDS-Shunts_1
Search Strategy:

| **#** | **Searches** | **Results** |
| --- | --- | --- |
| 1 | exp respiratory insufficiency/ | 63943 |
| 2 | Respiratory distress syndrome/ | 20797 |
| 3 | (respiratory adj2 insufficien*).tw,kf. | 9354 |
| 4 | (respiratory adj2 fail*).tw,kf. | 34680 |
| 5 | (respiratory adj2 distress*).tw,kf. | 44807 |
| 6 | ARDS*.tw,kf. | 14634 |
| 7 | AHRF*.tw,kf. | 195 |
| 8 | (CARDS or C-ARDS).tw,kf. | 10669 |
| 9 | COVID-19/ | 66643 |
| 10 | Coronavirus Infections/ | 44654 |
| 11 | Coronavirus/ | 4572 |
| 12 | Betacoronavirus/ | 33199 |
| 13 | SARS-CoV-2/ | 51893 |
| 14 | Covid*.tw,kf. | 104925 |
| 15 | (nCov or novel-CoV or 2019nCoV).tw,kf. | 1790 |
| 16 | (CoV-2 or CoV2 or sarscov2 or sarscov-2).tw,kf. | 38362 |
| 17 | Wuhan-virus*.tw,kf. | 18 |
| 18 | ((wuhan or hubei or huanan) and (severe-acute-respiratory or pneumonia*) and outbreak*).tw,kf. | 976 |
| 19 | or/1-18 [ARDS/COVID] | 266102 |
| 20 | Echocardiography, Transesophageal/ | 21268 |
| 21 | transpulmonary bubble.tw,kf. | 4 |
| 22 | agitated saline.tw,kf. | 350 |
| 23 | ((bubble or microbubble) adj3 (study or studies)).tw,kf. | 441 |
| 24 | (bubble adj3 echocardiogra*).tw,kf. | 125 |
| 25 | (Saline contrast adj3 (study or studies)).tw,kf. | 30 |
| 26 | TPBT*.tw,kf. | 8 |
| 27 | transthoracic echocardiogra*.tw,kf. | 14148 |
| 28 | ((transesophageal or transoesophageal) adj2 echocardiogra*).tw,kf. | 18799 |
| 29 | (vascular adj2 dilat*).tw,kf. | 1603 |
| 30 | ((intracardiac* or intra-cardiac* or cardiac*) adj3 shunt*).tw,kf. | 1727 |
| 31 | ((intrapulmonary* or intra-pulmonary* or pulmonary*) adj3 shunt*).tw,kf. | 4250 |
| 32 | or/20-31 [DIAGNOSING SHUNTS] | 47784 |
| 33 | 19 and 32 | 1190 |

Database(s): **Embase** 1974 to 2021 March 23

Search Title: ARDS-Shunts_2
Search Strategy:

| **#** | **Searches** | **Results** |
| --- | --- | --- |
| 1 | exp respiratory failure/ | 104935 |
| 2 | Respiratory distress syndrome/ | 14477 |
| 3 | Adult respiratory distress syndrome/ | 42416 |
| 4 | (respiratory adj2 insufficien*).tw,kw. | 11103 |
| 5 | (respiratory adj2 fail*).tw,kw. | 59497 |
| 6 | (respiratory adj2 distress*).tw,kw. | 65370 |
| 7 | ARDS*.tw,kw. | 24438 |
| 8 | AHRF*.tw,kw. | 387 |
| 9 | (CARDS or C-ARDS).tw,kw. | 15465 |
| 10 | exp Coronavirinae/ | 23440 |
| 11 | Coronavirus Infection/ | 13028 |
| 12 | SARS coronavirus/ | 6308 |
| 13 | Covid*.tw,kw. | 105002 |
| 14 | (nCov or novel-CoV or 2019nCoV).tw,kw. | 1823 |
| 15 | (CoV-2 or CoV2 or sarscov2 or sarscov-2).tw,kw. | 37348 |
| 16 | Wuhan-virus*.tw,kw. | 13 |
| 17 | ((wuhan or hubei or huanan) and (severe-acute-respiratory or pneumonia*) and outbreak*).tw,kw. | 1047 |
| 18 | or/1-17 [ARDS/COVID] | 336353 |
| 19 | Transesophageal echocardiography/ | 47413 |
| 20 | Contrast echocardiography/ | 4108 |
| 21 | Heart septum defect/ | 7685 |
| 22 | Pulmonary shunt/ | 1293 |
| 23 | transpulmonary bubble.tw,kw. | 5 |
| 24 | agitated saline.tw,kw. | 788 |
| 25 | ((bubble or microbubble) adj3 (study or studies)).tw,kw. | 866 |
| 26 | (bubble adj3 echocardiogra*).tw,kw. | 374 |
| 27 | (Saline contrast adj3 (study or studies)).tw,kw. | 61 |
| 28 | TPBT*.tw,kw. | 9 |
| 29 | transthoracic echocardiogra*.tw,kw. | 29407 |
| 30 | ((transesophageal or transoesophageal) adj2 echocardiogra*).tw,kw. | 28375 |
| 31 | (vascular adj2 dilat*).tw,kw. | 2316 |
| 32 | ((intracardiac* or intra-cardiac* or cardiac*) adj3 shunt*).tw,kw. | 2658 |
| 33 | ((intrapulmonary* or intra-pulmonary* or pulmonary*) adj3 shunt*).tw,kw. | 6029 |
| 34 | or/19-33 [DIAGNOSING SHUNTS] | 97883 |
| 35 | 18 and 34 | 3314 |

Database: Cochrane Library (Cochrane Reviews & Trials)
Search Name:	ARDS-Shunts_3

Date Run:	25/03/2021 10:21:08

Comment:

ID	Search	Hits

#1	[mh "respiratory insufficiency"]	2854

#2	[mh ^"Respiratory distress syndrome"]	1392

#3	(respiratory near/2 insufficien*):ti,ab,kw	2009

#4	(respiratory near/2 fail*):ti,ab,kw	4889

#5	(respiratory near/2 distress*):ti,ab,kw	6814

#6	ARDS*:ti,ab,kw	2034

#7	AHRF*:ti,ab,kw	86

#8	(CARDS or C-ARDS):ti,ab,kw	2260

#9	[mh ^"Covid-19"]	257

#10	[mh ^"Coronavirus infections"]	596

#11	[mh ^Coronavirus]	3

#12	[mh ^Betacoronavirus]	128

#13	[mh ^"SARS-CoV-2"]	204

#14	Covid*:ti,ab,kw	4497

#15	(nCov or "novel COV" or 2019nCoV):ti,ab,kw	141

#16	("COV-2" or COV2 or sarscov2 or "sarscov-2"):ti,ab,kw	1742

#17	"wuhan virus*":ti,ab,kw	0

#18	((wuhan or hubei or huanan) and ("severe acute respiratory" or pneumonia*) and outbreak*):ti,ab,kw	40

#19	{or #1-#18}	19342

#20	[mh ^"Echocardiography, transesophageal"]	418

#21	"transpulmonary bubble":ti,ab,kw	0

#22	"agitated saline":ti,ab,kw	26

#23	((bubble or microbubble) near/3 (study or studies)):ti,ab,kw	38

#24	(bubble near/3 echocardiogra*):ti,ab,kw	2

#25	("saline contrast" near/3 (study or studies)):ti,ab,kw	1

#26	TPBT*:ti,ab,kw	0

#27	"transthoracic echocardiogra*":ti,ab,kw	0

#28	((transesophageal or transoesophageal) near/2 echocardiogra*):ti,ab,kw	1101

#29	(vascular near/2 dilat*):ti,ab,kw	79

#30	((intracardiac* or "intra cardiac*" or cardiac*) near/3 shunt*):ti,ab,kw	66

#31	((intrapulmonary* or "intra pulmonary*" or pulmonary*) near/3 shunt*):ti,ab,kw	450

#32	{or #20-#31}	1720

#33	#19 and #32	90 (Only trials; no Cochrane reviews were available)

Database: DARE (Database of Abstracts for Reviews or Effects)
Search Strategy:
Results for: (respiratory insufficien* or respiratory fail* or respiratory distress* or ARDS* or AHRF* or CARDS or COVID* or SARS* or COV2* or COV-2* or nCov or novel cov* or corona* or pneumonia* outbreak* or severe acute respiratory) AND (transesophageal echocardiograp* or transoesophageal echocardiograp* or transthoracic echocardiograp* or transpulmonary bubble or agitated saline or bubble study or bubble studies or bubble echocardiograp* or saline contrast or TPBT* or vascular dilat* or shunt*) 23
Note: No filters were available to translate from DARE to EndNote, so they were entered by hand.
