## Supplemental Table 2 for "Acute respiratory distress syndrome and shunt detection with bubble studies: a systematic review and meta-analysis"

|  | Were the 2 groups similar and recruited from the same population? | Were the exposures measured similarly to assign people to both exposed and unexposed groups? | Was the exposure measured in a valid and reliable way? | Were the confounding factors identified? | Were strategies to deal with confounding factors stated? | Were the groups / participants free of the outcome at the start of the study (or at the moment of exposure)? | Were the outcome measured in a valid and reliable way? | Was the follow-up time reported sufficient to be long enough for outcome to occur? | Was follow up complete, and if not, were the reasons to loss to follow up described and explored? | Were strategies to address incomplete follow-up utilized? | Was appropriate statistical analysis used? | Overall appraisal |
| --- | --- | --- | --- | --- | --- | --- | --- | --- | --- | --- | --- | --- |
| **Observational cohort (9)** |  |  |  |  |  |  |  |  |  |  |  |  |
| Boissier 2015 | Yes | Yes | Yes | Yes | Unclear | Yes | Yes | Unclear | Yes | Unclear | Yes | Include |
| Legras 2015 | Yes | Yes | Yes | Unclear | Unclear | Yes | Yes | Yes | Yes | Unclear | Yes | Include |
| Lhertier 2013 | Yes | Yes | Yes | Unclear | Unclear | Yes | Yes | Yes | Yes | Yes | Yes | Include |
| Masi 2020 | NA | No | Unclear | No | No | Unclear | No | NA | NA | Unclear | NA | Exclude |
| Mekontso Dessap 2010 | Yes | Yes | Yes | Unclear | No | Yes | Yes | Yes | Yes | Unclear | Yes | Include |
| Mekontso Dessap 2011 | NA | No | Unclear | No | Unclear | Unclear | No | NA | NA | Unclear | NA | Exclude |
| Salazar-Orellana 2021 | Yes | Yes | Yes | Yes | Unclear | Yes | Yes | Unclear | Unclear | Unclear | Yes | Include |
| Vavlitou 2010 | Yes | Yes | Yes | Unclear | Unclear | Yes | No | NA | NA | Unclear | NA | Exclude |
| Vedrienne 1995 | Unclear | Yes | Yes | No | No | Unclear | Unclear | Yes | Yes | Unclear | Yes | Include |
| **Cross-sectional pilot study (1)** |  |  |  |  |  |  |  |  |  |  |  |  |
| Reynolds 2020 | NA | No | Unclear | Unclear | No | Unclear | No | NA | NA | Unclear | NA | Exclude |

Supplemental Table 2. Composite risk of bias assessment utilizing the Joanna-Briggs Institute (JBI) critical appraisal tool for cohort and case control studies.
