## Supplemental Table 3 for "Acute respiratory distress syndrome and shunt detection with bubble studies: a systematic review and meta-analysis"

**Reference:**	Representative ness of exposed

cohort (/1)

### Observational cohort (9)

Selection of non-exposed cohort

(/1)

Ascertainment of exposure (/1)

Demonstration outcome interest not present at initiation of study (/1)

Comparability of cohorts (/2)

Assessment of outcome (/1)

Was follow-up long enough for outcome to occur?

(/1)

Was follow- up adequate (/1)

### Total score ( /9)

**Overall Risk of Bias**

Boissier 2015	1	1	1	1	0	1	1	1	**7	Poor**

Legras 2015	1	1	1	1	1	1	1	1	**8	Good**

Lhertier 2013	1	1	1	1	1	1	1	1	**8	Good**

Masi 2020	1	1	1	1	0	0	0	0	**4	Poor**

Mekontso Dessap 2010	1	1	1	1	1	1	1	1	**8	Good**

Mekontso Dessap 2011	1	1	1	1	1	0	0	0	**5	Poor**

Salazar-Orellana 2021	1	1	1	1	1	1	0	1	**7	Good**

Vavlitou 2010	1	1	1	1	1	0	0	0	**5	Poor**

Vedrienne 1995	1	1	1	0	1	0	1	1	**6	Fair**

### Cross-sectional pilot study (1)

| Reynolds 2020 | 1 | 1 | 1 | 1 | 1 | 0 | 0 | 0 | **5** | **Poor** |
| --- | --- | --- | --- | --- | --- | --- | --- | --- | --- | --- |

**Supplemental Table 3**. Composite risk of bias assessment utilizing the Newcastle-Ottawa Risk of Bias Scale

|  | Representativeness of exposed cohort (/1) | Representativeness of non-exposed cohort (/1) | Ascertainment of exposure (/1) | Demonstration outcome of interest not present at initiation of study (/1) | Comparability of cohorts (/2) | Assessment of outcome (/1) | Was follow-up long enough for outcome to occur? (/1) | Was follow-up adequate? (/1) | Total score (/9) | Overall Risk of Bias |
| --- | --- | --- | --- | --- | --- | --- | --- | --- | --- | --- |
| Observational cohort (9) |  |  |  |  |  |  |  |  |  |  |
| Boissier 2015 | 1 | 1 | 1 | 1 | 0 | 1 | 1 | 1 | **7** | **Poor** |
| Legras 2015 | 1 | 1 | 1 | 1 | 1 | 1 | 1 | 1 | **8** | **Good** |
| Lhertier 2013 | 1 | 1 | 1 | 1 | 1 | 1 | 1 | 1 | **8** | **Good** |
| Masi 2020 | 1 | 1 | 1 | 1 | 0 | 0 | 0 | 0 | **4** | **Poor** |
| Mekontso Dessap 2010 | 1 | 1 | 1 | 1 | 1 | 1 | 1 | 1 | **8** | **Good** |
| Mekontso Dessap 2011 | 1 | 1 | 1 | 1 | 1 | 0 | 0 | 0 | **5** | **Poor** |
| Salazar-Orellana 2021 | 1 | 1 | 1 | 1 | 1 | 1 | 0 | 1 | **7** | **Good** |
| Vavlitou 2010 | 1 | 1 | 1 | 1 | 1 | 0 | 0 | 0 | **5** | **Poor** |
| Vedrienne 1995 | 1 | 1 | 1 | 0 | 1 | 0 | 1 | 1 | **6** | **Fair** |
| Cross-sectional pilot study (1) |  |  |  |  |  |  |  |  |  |  |
| Reynolds 2020 | 1 | 1 | 1 | 1 | 1 | 0 | 0 | 0 | **5** | **Poor** |
