## Supplementary figures and images for "Acute respiratory distress syndrome and shunt detection with bubble studies: a systematic review and meta-analysis"

### Supplemental Figure 1

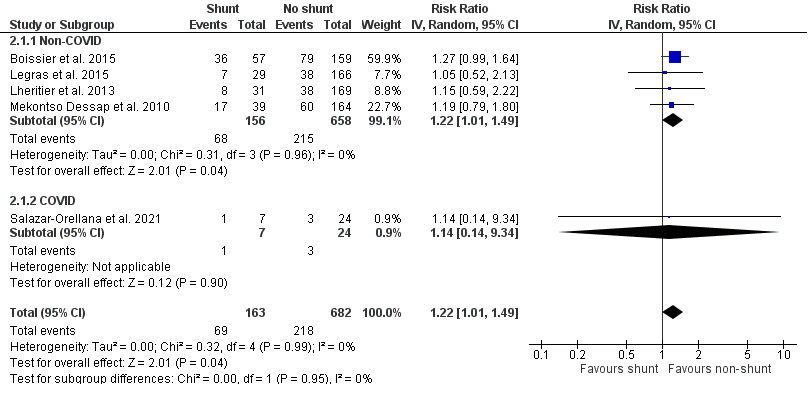

### Supplemental Figure 2

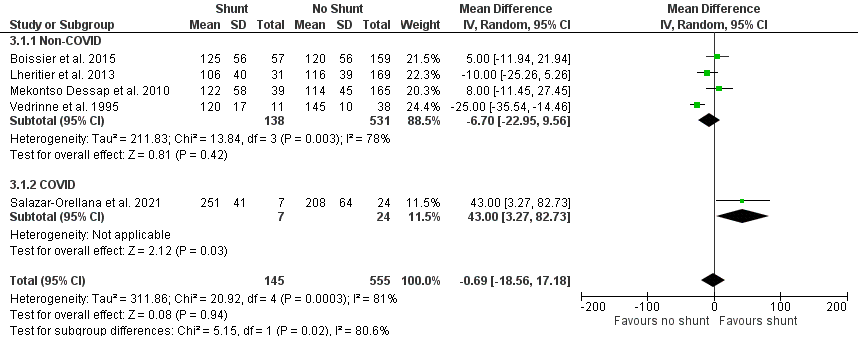
